# Immunogenicity and safety of an Omicron XBB.1.16 adapted vaccine for COVID-19: Interim results from a randomized, controlled, non-inferiority clinical trial

**DOI:** 10.1101/2024.04.19.24306064

**Authors:** María Jesús López Fernandez, Silvia Narejos, Antoni Castro, José María Echave-Sustaeta, María José Forner, Eunate Arana-Arri, Josep Molto, Laia Bernad, Raúl Pérez-Caballero, Julia G Prado, Dàlia Raïch-Regué, Rytis Boreika, Nuria Izquierdo-Useros, Julià Blanco, Joan Puig-Barberà, Silvina Natalini Martínez

**Affiliations:** Preventive Medicine Unit, Hospital Regional Universitario de Málaga, Málaga, Spain; Centro de Atención Primaria Centelles, Centelles, Spain; Hospital Universitari de Girona Doctor Josep Trueta, Girona, Spain; Hospital Universitario Quirón Salud Madrid, Madrid, Spain; Hospital Clínico Universitario Valencia, Valencia, Spain; Unidad de Coordinación Científica, Biocruces Bizkaia, Osakidetza, Barakaldo, Spain; Centro de Investigación Biomédica en Red-Enfermedades Infecciosas (CIBERINFEC), Instituto de Salud Carlos III, Madrid, Spain; Fundació Lluita contra les Infeccions, Department of Infectious Diseases, Hospital Universitari Germans Trias i Pujol, Badalona, Spain; IrsiCaixa, Can Ruti Campus, Badalona, Spain; Institut de Recerca Germans Trias i Pujol (IGTP), Badalona, Spain; Càtedra de Malalties Infeccioses i Immunitat, Facultat de Medicina, Universitat de Vic-Universitat Central de Catalunya (UVic-UCC), Barcelona, Spain; Área de Investigación en Vacunas, Fundació per al Foment de la Investigació Sanitària i Biomèdica de la Comunitat Valenciana (FISABIO), Valencia, Spain; Unidad de Investigación de Vacunas, Instituto de Investigación Sanitaria HM, Madrid, Spain

**Keywords:** JN.1, XBB.1.16, adapted vaccine, SARS-COV-2 vaccine, adjuvanted protein vaccine, booster vaccine, COVID-19

## Abstract

**Background:** Global COVID-19 vaccination adapts to protect populations from emerging variants. This communication presents interim findings from the new Omicron XBB adapted PHH-1V81 vaccine compared to a XBB adapted mRNA vaccine against XBB and JN.1 SARS-CoV-2 strains.

**Methods:** In a Phase IIb/III pivotal trial, adults previously vaccinated with a primary scheme and at least one booster dose of an EU-approved mRNA vaccine randomly received either PHH-1V81 or BNT162b2 XBB.1.5 vaccine booster as a single dose. The primary efficacy endpoint assessed neutralisation titers against the Omicron XBB.1.16 variant at day 14. Secondary endpoints evaluated neutralization titers and cellular immunity against different variants. Safety endpoints comprised solicited reactions up to day 7 post-vaccination and serious adverse events until the cut-off date of the interim analysis. Changes in humoral responses were reported as GMT and GMFR assessed by PBNA or VNA.

**Results:** At the cut-off date, immunogenicity assessments included 599 participants. Both boosters elicited neutralizing antibodies against XBB.1.5, XBB.1.16 and JN.1 with PHH-1V81 inducing a higher response for all variants. PHH-1V8 booster triggers a superior neutralizing antibodies response against XBBs variants compared to the mRNA vaccine. Subgroup analysis consistently revealed higher neutralizing antibody responses with PHH-1V81 across age groups, number of prior vaccination shots, and SARS-CoV-2 infection history. Safety analysis involved 607 participants at the day 14 visit, revealing favourable safety profiles without any serious vaccine-related adverse events at cut-off date of the interim analysis (12^th^ December 2023).

**Conclusions:** PHH-1V81 demonstrates superiority on humoral immunogenicity compared to mRNA vaccine agains XBB variants and non-inferiority against JN.1 with favourable safety profile and lower reactogenicity, confirming its potential as vaccine candidate.

## 1. Introduction

The ongoing coronavirus disease 2019 (COVID-19) pandemic has spurred extensive vaccination efforts worldwide to combat the severe acute respiratory syndrome coronavirus 2 (SARS-CoV-2) and its variants.^1–3^ While current vaccines have shown effectiveness, their protection diminishes over time, particularly against emerging variants.^4–6^ This highlights the crucial need to adapt vaccine compositions and develop updated strategies to sustain immunity.^7^

Thus, with the emergence of Omicron XBB variants, regulatory and public health bodies such as the World Health Organization (WHO) Technical Advisory Group on COVID-19 Vaccine Composition (TAG-CO-VAC), the European Centre for Disease Prevention and Control (ECDC) and the European Medicines Agency (EMA) recommended updates to specifically target viral lineages for primary or booster vaccinations.^8,9^ Responding to these recommendations, mRNA and protein-based vaccines targeting Omicron XBB have gained approval and are widely used.^9–13^ The emergence of the JN.1 variant ongoing pandemic challenges, highlighting the critical need for vaccine advancement to address evolving variants effectively.^14,15^

PHH-1V, a bivalent protein-based adjuvanted vaccine, has emerged as a promising booster.^16^ Phase IIb trials demonstrated its efficacy in generating neutralizing antibodies against various SARS-CoV-2 variants, including Omicron XBB.^16–18^ Long-term analysis revealed sustained immune responses, even in high-risk populations regardless of prior infection or primary vaccine type.^19^ Additionally, PHH-1V induces a robust T-cell response.^17,18^

Further, recipients of PHH-1V reported fewer adverse events (AE) than mRNA vaccine recipients, with comparable breakthrough non-severe COVID-19 rates. As a recombinant protein-based vaccine, PHH-1V also may offer advantages, including high productivity, stability, and suitability for immunocompromised individuals, positioning it as a promising candidate in the battle against COVID-19.^20^ A new adapted XBB vaccine, PHH-1V81, based on an XBB.1.16 fusion RBD homodimer has been developed in response to SARS-CoV-2 evolution.

Here, interim findings from the HIPRA-HH-14 clinical study are reported. The HIPRA-HH-14 clinical study is a Phase IIb/III pivotal non-inferiority trial evaluating the immunogenicity and safety of the XBB.1.16 monovalent adapted vaccine, PHH-1V81, as a booster. It examines immunogenicity changes against various SARS-CoV-2 variants, including Omicron XBB.1.16, Omicron XBB.1.5 and emerging variants such as JN.1 in a subset of participants.

## 2. Material and methods

### Study design

This HIPRA-HH-14 Phase IIb/III pivotal trial is a double-blind, randomized, active-controlled, multi-centre, non-inferiority clinical study assessing the efficacy, safety and tolerability of PHH-1V81 XBB.1.16 adapted booster vaccine, targeting the Omicron XBB.1.16 variant of SARS-CoV-2 compared to a mRNA XBB.1.5 adapted vaccine. Participants had previously received two doses of an EU-approved mRNA vaccine and at least one booster dose.

The primary efficacy endpoint is neutralization titers against Omicron XBB.1.16 variant at day 14. Secondary efficacy endpoints include neutralization titers against Omicron XBB.1.16 variant at days 91 and 182, and against the Wuhan, Omicron BA.1, Omicron XBB.1.5 strains at days 14, 91 and 182. Safety endpoints comprise solicited systemic and local reactions up to day 7 post-vaccination, unsolicited local and systemic AE through Day 28 after vaccination, AE of special interest (AESI) through the end of study, related medically attended AE through the end of study and serious AE (SAE) throughout the study period.

The planned interim analysis reports the results comparing the immunogenicity and safety of PHH-1V81 (HIPRA) with BNT162b2 XBB.1.5 (Pfizer-BioNTech) at baseline and day 14, including solicited AE at day 7 post-vaccination for participants with completed day 14 visit and SAE until the cut-off date of the interim analysis (12^th^ December 2023).

Humoral response was evaluated by measuring the inhibitory concentration 50 (IC50) using a pseudovirion-based neutralization assay (PBNA) against Omicron XBB.1.16 and Omicron XBB.1.5 variants, reported as geometric mean titer (GMT) and geometric mean fold rises (GMFR) for adjusted treatment.

Cellular responses were assessed through interferon-γ (IFN-γ) ELISpot assay as an exploratory endpoint. With this purpose, peripheral blood mononuclear cells (PBMC) were re-stimulated in vitro with 6 SARS-CoV-2 RBD peptides’ pools (Wuhan, Omicron BA.1, Omicron XBB.1.5, Omicron XBB.1.16, Omicron BA.2.86 and Omicron JN.1 variants) at baseline and day 14 in a subset of participants, to determine the percentage of antigen-specific IFN-γ producing T-cells.^18^

In addition to planned immunogenicity assessments a virus neutralization assay (VNA) was conducted in a random subset of serum samples to compare humoral immune response between vaccine arms against the Omicron JN.1.^21^ ^22^

The trial was conducted in accordance with the Declaration of Helsinki, the Good Clinical Practice guidelines, and national regulations^23–25^. The study protocol was reviewed and approved by the Spanish Agency of Medicines and Medical Devices (AEMPS) and by Independent Ethics Committee from HM Hospitales (23.10.2249-GHM).

### Participants

The trial enrolled adults aged 18 or older who provided informed consent, previously received a primary scheme of two doses and at least one booster dose of an EU-approved mRNA vaccine with last dose at least six months prior inclusion and tested negative for acute SARS-CoV-2 infection on day 0. Participants with stable chronic diseases were eligible. Participants with a previous SARS-CoV-2 infection must have been diagnosed at least 6 months before day 0. Full selection criteria are provided in the Supplementary Appendix. The trial is being conducted at ten clinical sites in Spain, with enrolment starting on November 15th, 2023. Rapid recruitment led to trial closure on November 29th, 2023.

### Randomization and treatment allocation

Participants were randomly assigned to treatment arms in a 2:1 ratio to receive either a booster dose of PHH-1V81 (HIPRA XBB adapted vaccine, n= 408) or a booster dose of BNT162b2 Omicron XBB.1.5 (Comirnaty® Omicron XBB.1.5, Pfizer-BioNTech adapted vaccine, n=204).

All participants received their respective booster dose on day 0 and were closely observed for 15 minutes after vaccination on-site.

### Sample size

In the HIPRA-HH-14 trial, sample size determination aimed to confirm PHH-1V81’s non-inferiority to the comparator vaccine in inducing neutralizing antibody titers against Omicron XBB variants. The success criterion was defined as the upper bound of the 95% confidence interval (CI) around the GMT ratio BNT162b2 XBB.1.5: PHH-1V81 should lie below 1.50. With a 2:1 randomization ratio, group sizes of 366 and 183 ensured 90% power at a one-sided 2.5% significance level. This resulted in 612 randomized subjects, with 408 in the PHH-1V81 arm, providing ample power for AE detection.

### Statistical analysis

Descriptive analyses compared time points and treatment arms for each SARS-CoV-2 variant. Categorical variables were presented as cases and percentages, while continuous variables included non-missing observations, mean (or geometric mean), standard deviation (or geometric standard deviation), median, interquartile range, minimum, and maximum, without imputation for missing data. Efficacy analyses followed predefined hypotheses for noninferiority, with the upper bound of the 95% CI determining claim validation. GMT and GMFR adjusted treatment for immunogenicity endpoints were estimated using Mixed Models for Repeated Measures (MMRM), while T-cell data were analysed with mixed effects models. Values below the lower limit of quantification (LLOQ) were imputed as LLOQ, and PBNA values exceeding 20480 were reanalysed.

### Results

Due to the rapid recruitment of the trial, a total of 905 subjects constituted the Intention-to-Treat (ITT) population out of 913 screened (293 subjects more than the calculated sample size). Among them, 603 were allocated to the PHH-1V81 vaccine arm and 302 to the BNT162b2 XBB.1.5 vaccine arm. As per the electronic case report form (eCRF) data collected up to December 12th, 2023, vaccine administration was confirmed for 800 subjects (536 in PHH-1V81; 264 in BNT162b2 XBB.1.5), forming the safety population for interim analysis. However, only 607 subjects (409 in PHH-1V81; 198 in BNT162b2 XBB.1.5) had completed the day 14 visit and information on solicited AE was available for the safety interim analysis. Of these, immunogenicity data at both baseline and day 14 visits was available for 599 participants (66.2% of ITT, 97.9% of target sample size), which were included in the modified Intention-to-Treat (mITT) population for immunogenicity assessments, as specified in the clinical study protocol. Of these, 406 received the PHH-1V81 booster vaccine and 193 received the BNT162b2 XBB.1.5 booster vaccine. No premature discontinuations occurred in the study as of the cut-off date ***(Figure 1)*.**

**Figure 1.**
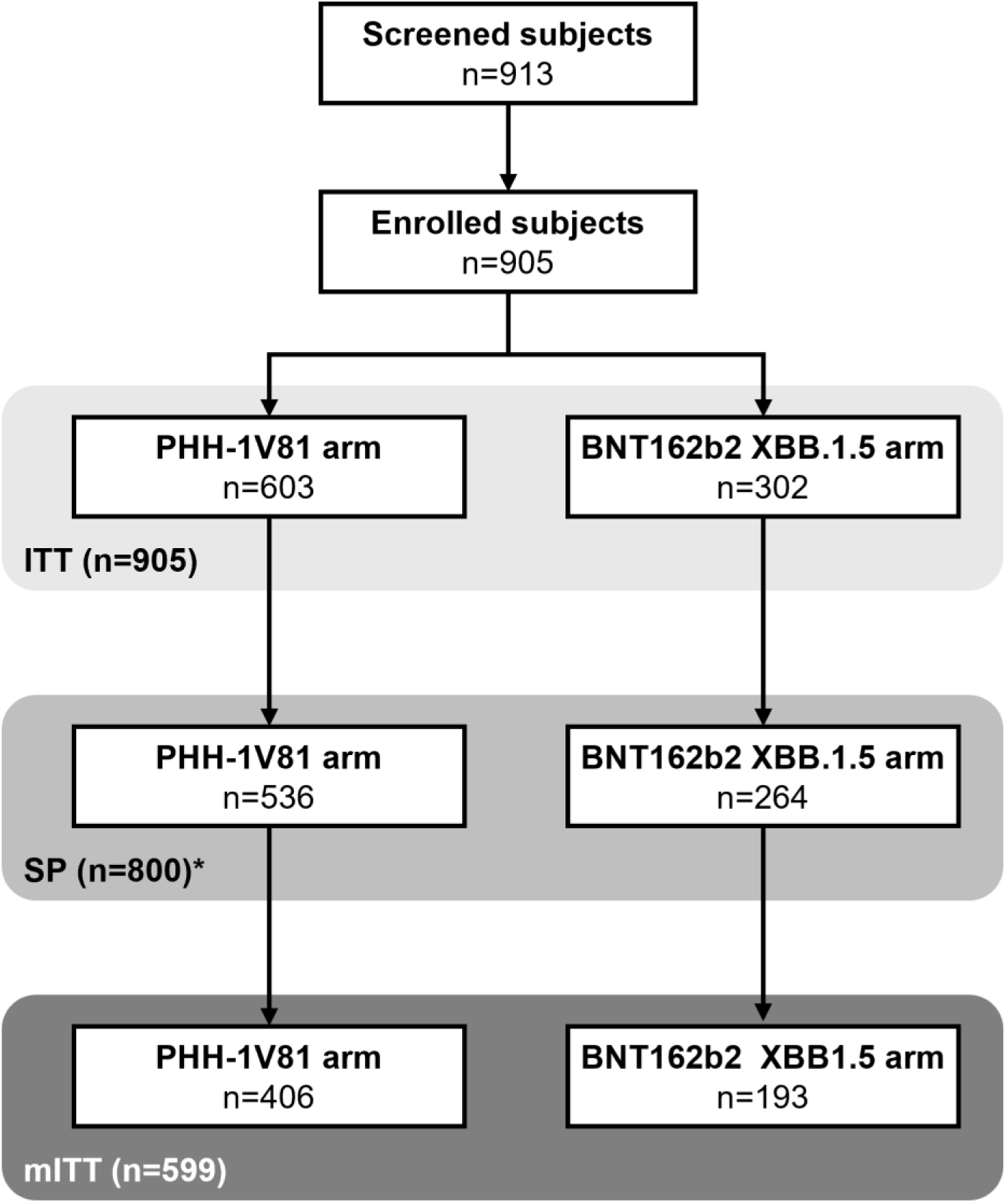
Participants disposition of HIPRA-HH14. The Enrolled Population (EP) is defined as all subjects who signed the Informed Consent Form. The Intention-to-treat Population (ITT) is defined as all subjects of the EP who are randomly assigned to treatment, regardless of the subject’s treatment status in the study. The Safety Population (SP) set is defined as all randomised subjects who received the study drug. * 607 subjects (409 in PHH-1V81 arm and 198 in the BNT162b2 XBB.1.5 arm) had completed Day 14 visit and information on solicited adverse events was available at the cut-off date set on 12th December 2023. The Modified ITT Population (mITT) is defined as all participants in the ITT who met the inclusion/exclusion criteria and received a dose of study drug, whose Baseline and Day 14 were available and did not test positive for COVID-19 within 14 days of receiving study drug.

### Baseline characteristics

The median age of participants was 45 years (range: 18 to 88 years) with similar age ranges in both vaccine arms. Most subjects were 18 to 59 years old (87.0%), female (59.3%), white (93.8%), who had received 3 (66.9%) or 4 (33.0%) previous vaccination doses. Demographic features were generally balanced between the vaccine arms ***(Table S1 in the Supplementary Appendix)*.**

### PHH-1V81 immunogenicity against Omicron XBB.1.16 and Omicron XBB.1.5 variants

The booster immunization with both vaccines induces a remarkable increase in neutralizing antibodies 14 days after vaccination compared to baseline against the Omicron XBB.1.16 and Omicron XBB.1.5 variants ***(Figure 2 and Table 1).*** The GMT (95% CI) for adjusted treatment against Omicron XBB.1.16 increased from 152.46 (134.72, 172.54) at baseline to 1946.38 (1708.44, 2217.46) at day 14 after the PHH-1V81 booster, and from 161.57 (136.40, 191.37) at baseline to 1512.21 (1261.72, 1812.44) at day 14 after the BNT162b2 XBB.1.5 booster. The GMT for adjusted treatment against Omicron XBB.1.5 increased from 151.93 (134.89, 171.13) at baseline to 1888.89 (1676.99, 2127.57) at day 14 after the PHH-1V81 booster, and from 167.89 (142.04, 198.44) at baseline to 1486.03 (1257.25, 1756.45) at day 14 after the BNT162b2 XBB.1.5 booster.

**Figure 2.**
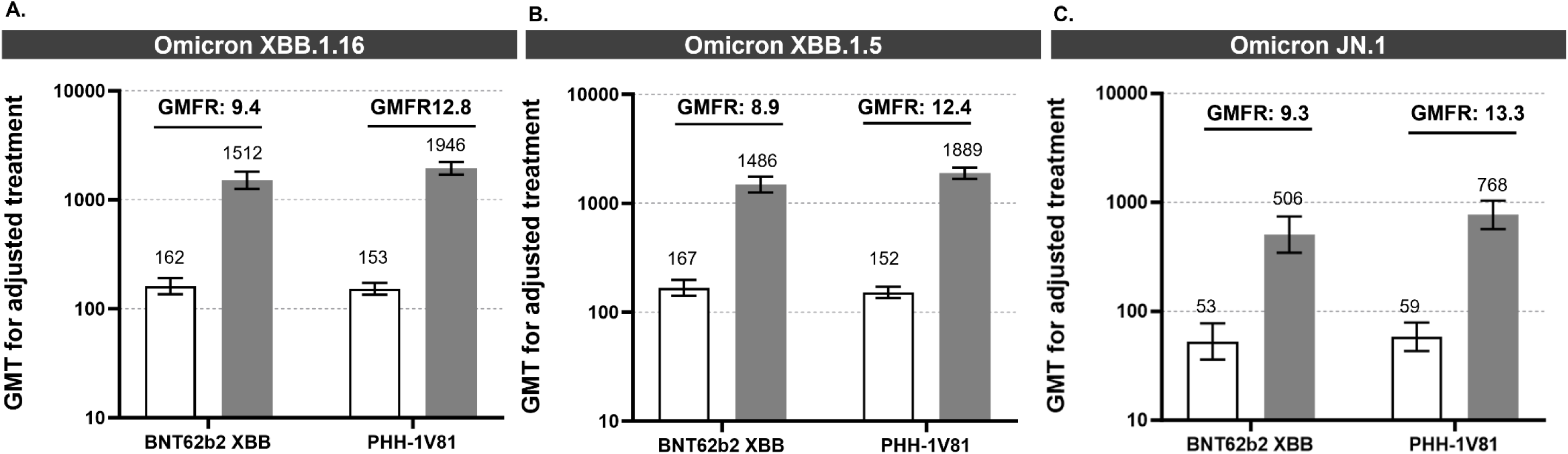
Humoral response against SARS-CoV-2 variants Omicron XBB.1.16, Omicron XBB.1.5 and Omicron JN.1 induced by BNT62n2 XBB and PHH-1V81 at Day 14. **A.**) GMT for adjusted treatment against Omicron XBB.1.16 variant for each vaccinated group (PHH-1V81 group; n= 406 and BNT62b2 XBB; n=193) by PBNA; (**B.**) GMT for adjusted treatment against Omicron XBB.1.5 variant for each vaccinated group (PHH-1V81 group; n= 406 and BNT62b2 XBB; n=193) by PBNA; (**C**.) GMT for adjusted treatment against Omicron JN.1 variant for a subset of n=100 participants (n= 65 with PHH-1V81 and n=35 with BNT62b2 XBB1.5) by VNA. Graphics A-C represent Mean GMT with 95% CI at baseline (with bar) and 14 days after booster (dark grey bar); upper numbers represent mean GMFR at day 14 from baseline. CI: confidence interval; GMT: Geometric Mean Titer; GMFR: Geometric Mean Fold Rise.

This increase in neutralizing antibodies against both variants is reflected in a GMFR (95% CI) for adjusted treatment. The GMFR at day 14 against Omicron XBB.1.16 was 12.76 (11.01, 14.78) for the PHH-1V81 booster and 9.42 (7.61, 11.66) for the BNT162b2 XBB.1.5 booster. Similarly, the GMFR at day 14 against Omicron XBB.1.5 was 12.42 (10.62, 14.51) for the PHH-1V81 booster and 8.88 (7.20, 10.94) for the BNT162b2 XBB.1.5 booster.

Comparisons between vaccine arms revealed higher neutralizing antibody levels after boosting with PHH-1V81 compared to the BNT162b2 XBB.1.5 booster against both SARS-CoV-2 variants analysed, Omicron XBB.1.16 and Omicron XBB.1.5, showing significant differences in GMT at day 14 post-booster with a GMT ratio between vaccine arms at day 14 of 0.78 (95% CI: 0.63, 0.96; p<0.05) against Omicron XBB.1.16 and 0.79 (95% CI: 0.64, 0.96; p<0.05) against Omicron XBB.1.5. However, no differences were found at baseline. Additionally, significant differences in GMFR at day 14 were observed between vaccination arms ***(Figure 3)*.**

**Figure 3.**
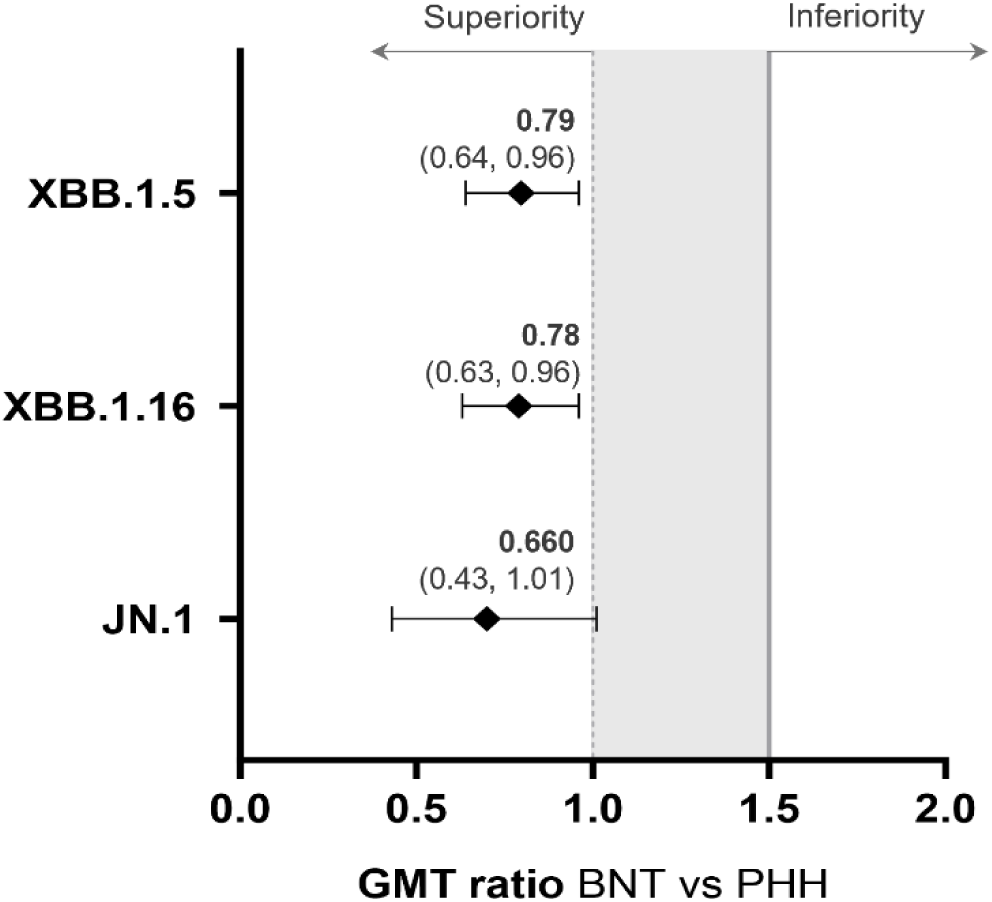
Comparison of the humoral responses elicited by BNT62b2 XBB and PHH-1V81 at Day 14 against SARS-CoV-2 variants Omicron XBB.1.16, Omicron XBB.1.5 and Omicron JN.1. Forest plot for GMT ratio (95% CI) BNT62b2 XBB.1.5 vs PHH-1V81 at day 14. The solid line indicates the non-inferiority limit of the trial (NIm = 1.5) and the dashed line indicates the superiority limit (GMT ratio = 1.0). BNT: BNT62b2 XBB1.5 vaccine; CI: confidence interval; GMT: Geometric Mean Titer; GMFR: Geometric Mean Fold Rise; PHH: PHH-1V81 vaccine

In the PHH-1V81 vaccine arm, 77.6% (95% CI: 73.2, 81.6%) and 76.4% (71.91, 80.41%) of subjects showed a ≥4-fold rise in neutralizing antibody titers against Omicron XBB.1.16 and Omicron XBB.1.5, respectively. In contrast, the BNT162b2 XBB.1.5 vaccine arm showed 71.0% (64.0, 77.3%) and 70.5% (63.49, 76.80%) of subjects demonstrating a ≥4-fold rise in neutralizing antibody titers against the respective variants.

### PHH-1V81 immunogenicity against Omicron XBB.1.16 and Omicron XBB.1.5 variants by participant subgroups

Subgroup analyses on neutralizing antibody titers against the Omicron XBB.1.16 and Omicron XBB.1.5 variants at day 14 revealed that, in participants aged 60 and above, the PHH-1V81 booster generated higher neutralizing antibody levels against both variants compared to BNT162b2 XBB.1.5. Similarly, individuals with and without prior SARS-CoV-2 infection exhibited increased neutralizing antibody levels following PHH-1V81 vaccination, surpassing levels induced by BNT162b2 XBB.1.5. Further, participants with three or more prior COVID-19 vaccine doses also showed numerically superior neutralizing antibody responses to Omicron XBB.1.16 and XBB.1.5 following PHH-1V81 boosting compared to BNT162b2 XBB.1.5 ***(Figures S1-S3 and Tables S3-S5 in the Supplementary Appendix)*.**

### PHH-1V81 immunogenicity against Omicron JN.1 variant

The booster dose with PHH-1V81 resulted in a significant increase in neutralizing antibody titers against Omicron JN.1 variant by VNA at day 14 after the booster, compared to baseline titers for both vaccination arms. Neutralizing antibody titers against JN.1 by VNA at baseline and day 14 post-boost were analysed in a random subset of mITT population of 100 participants (65 in PHH-1V81; 35 in BNT62b2). PHH-1V81 boost showed numerically higher neutralizing antibodies at day 14 against Omicron JN.1 compared to BNT62b2 Omicron XBB.1.5 with a GMT (95% CI) for adjusted treatment of 768.44 (568.96, 1037.86) and 505.88 (344.70, 742.43), respectively, and a GMT ratio of 0.66 (0.43, 1.01), indicating non-inferiority of the PHH-1V81 booster against the JN.1 variant. The GMFR (95% CI) at day 14 was 13.34 (8.84, 20.12) for PHH-1V81 and 9.27 (5.70, 15.07) for BNT62b2 Omicron XBB.1.5 ***(Figure 2 and Table S6 in the Supplementary Appendix)*.**

### PPH-1V81 cellular immunogenicity

SARS-CoV-2 specific T-cell responses after booster were evaluated in a subset of 40 participants at the cut-off date (27 in PPH-1V81; 13 in BNT162b2 XBB.1.5) by ELISpot from PBMC at baseline, and at 14 days. The participants included had a median age of 44 years (range: 23 to 79), with 4 individuals aged 60 years or older (3 in PHH-1V81; 1 in BNT162b2 XBB.1.5).

Both vaccines significantly increased the number of antigen-specific IFN-γ^+^ T-cells in response to *in vitro* PBMC re-stimulation with receptor-binding domain (RBD) peptide pools from, Omicron XBB.1.5, Omicron XBB.1.16, and Omicron JN.1 variants at day 14 post-booster compared to baseline. There were no significant differences in IFN-γ^+^ spot forming cells between the vaccine arms (***Figure 4 and Table S7 in the Supplementary Appendix)*.**

**Figure 4.**
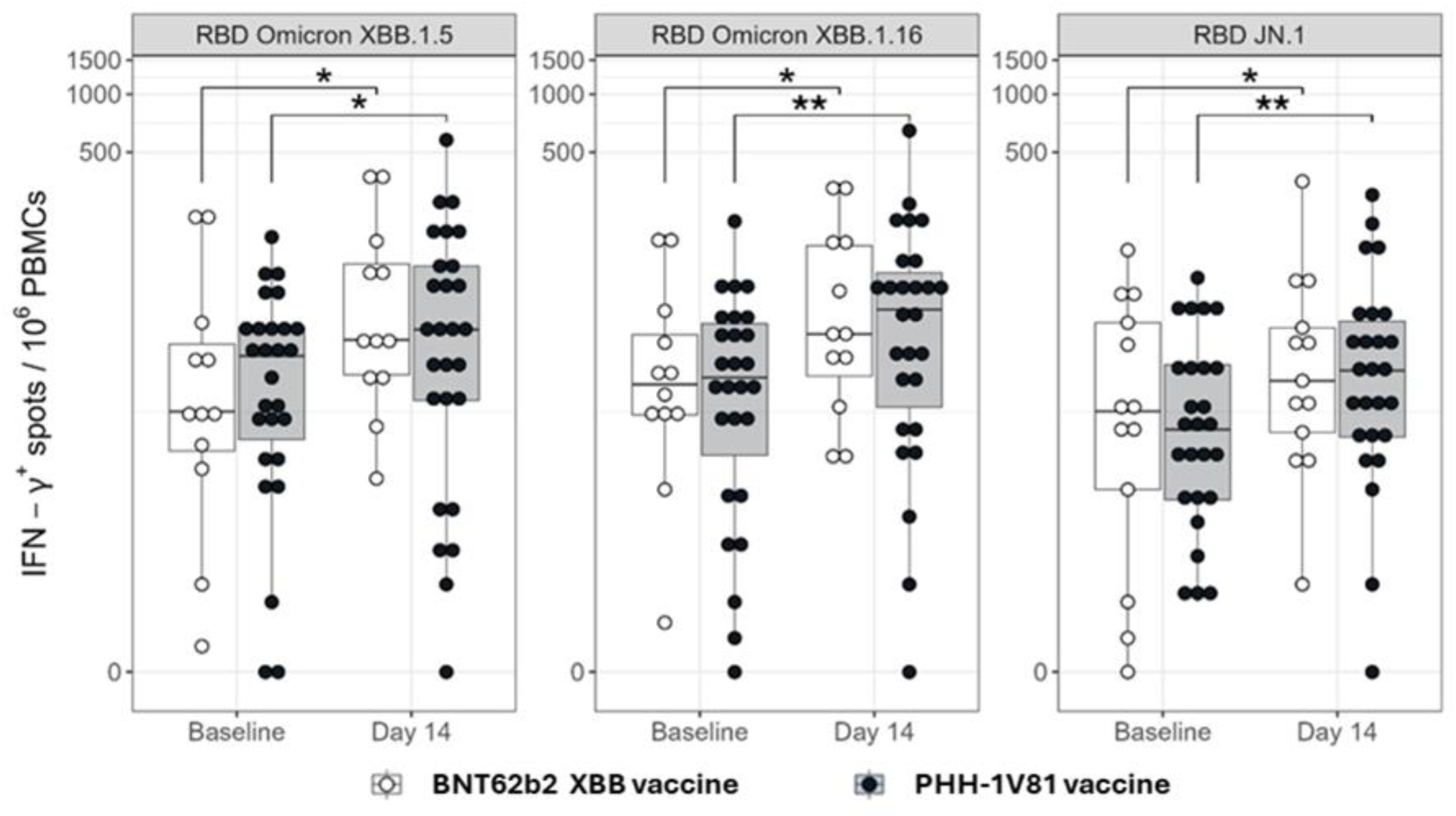
IFN-γ producing T cells upon PBMC re-stimulation with SARS-CoV-2 derived peptide pools, by vaccine arm. Frequencies of IFN-γ responses determined by ELISpot assay in PBMC from subgroup of participants immunised with PHH-1V81 (n= 27) and BNT162b2 XBB.1.5 (n=13). PBMC were isolated before the boost immunization (Baseline) and 2 weeks (D14) after boost with PHH-1V81 and BNT162b2 XBB.1.5 vaccines, stimulated with RBD Omicron XBB.1.5, Omicron XBB.1.16 and Omicron JN.1 peptides’ pools, and analysed by IFN-γ-specific ELISpot assay. Within group contrasts have been displayed in the plots, comparing the extent of IFN-γ+ response between timepoints in each treatment arm. Statistically significant differences between Baseline and Day 14 are shown in blue colour as * p< 0.01; ** p< 0.001; *** p<0.0001. IFN-γ: interferon- -γ; PBMC: peripheral blood mononuclear cells whole; RBD: receptor binding domain

### Safety and tolerability results

Among the 607 participants who completed the day 14 visit and provided information on solicited AE by December 12th, 2023, 163 reported no AE, with 118 in the PHH-1V81 arm and 45 in the BNT162b2 XBB.1.5 arm.

Most solicited local AE were mild in intensity, with 456 events in 210 subjects (51.3%), and 266 events in 118 subjects (59.6%) in the PHH-1V81 and BNT162b2 XBB.1.5 arms, respectively. Only 24 (5.9%) and 11 (5.6%) subjects in the PHH-1V81 and BNT162b2 XBB.1.5 arms, respectively, reported solicited local events of moderate intensity. One subject (0.2%) in the PHH-1V81 vaccine arm reported a severe local solicited event. Injection site pain, tenderness, discomfort was the most common solicited local AE, with an incidence of 59.1% and 51.3% in the BNT162b2 XBB.1.5 and PHH1V81 arms, respectively (***Figure 5)***. No serious AEs related to the study vaccines were reported.

**Figure 5.**
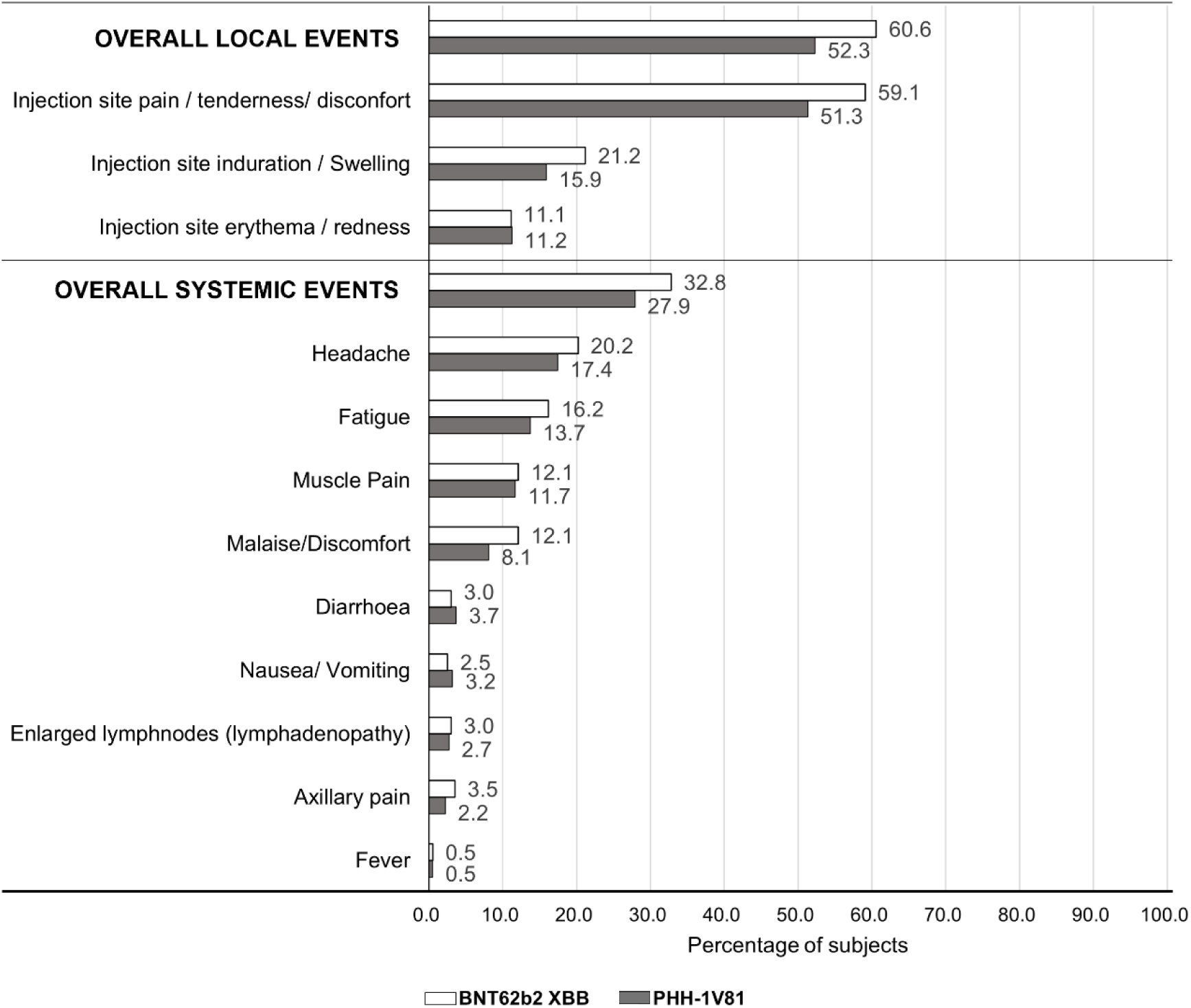
Percentage of subjects with solicited local and systemic adverse events through Day 7, by vaccine arm. Solicited local adverse events and solicited systemic adverse events were reported MedDRA. PT from Day 0 through Day 7 for the safety population with available data at the cut-off date. Data are shown as the percentage of subjects in relation to the safety population (n=607; n=409 in PHH-1V81 arm and n=198 in BNT162b2 XBB.1.5 arm). If a subject experienced more than one event, the subject is counted once for each type of event. PTs are ordered in decreasing frequency of the total number of subjects with each adverse event in PHH-1V81 group. n, the number of subjects in the population; PT, preferred term.

Regarding solicited systemic AE, most were also mild (grade 1), with 369 events in 167 subjects (27.5%), slightly lower in the PHH-1V81 arm (26.2%) compared to the BNT162b2 XBB.1.5 arm (30.3%). Moderate (grade 2) AE occurred in 63 events among 38 subjects (6.3%), with a lower incidence in the PHH-1V81 arm (4.6%) compared to the BNT162b2 XBB.1.5 arm (9.6%). Severe (grade 3) systemic AE were rare, affecting only 4 subjects (2 (0.5%) in PHH-1V81; 2 (1.0%) in BNT162b2 XBB.1.5). The most frequent systemic AEs were headache, fatigue, and muscle pain, with higher incidences in the BNT162b2 XBB.1.5 arm (***Figure 5)***.

Overall, the frequency of solicited local and systemic AE was higher in the BNT162b2 XBB.1.5 arm (32.8%) compared to the PHH-1V81 arm (27.9%) ***(Table S8 in the Supplementary Appendix)***. Additionally, 19.4% of PHH-1V81 recipients reported no AE, compared to 7.4% in the BNT162b2 XBB.1.5 arm.

## 3. Discussion

The interim analysis, conducted before study completion, compared the immunogenicity of PHH-1V81 with BNT162b2 XBB.1.5 against Omicron XBB 1.16. Secondary endpoints and additional analysis evaluated neutralization titers against XBB.1.5 and Omicron JN.1. Results revealed that 14 days post-vaccination, the immune response against XBB.1.16 with PHH-1V81 booster vaccine was consistently not only non-inferior but also superior to that with the BNT162b2 XBB.1.5 booster vaccine. Additionally, PHH-1V81 significantly increased neutralizing antibody titers against XBB.1.5 and JN.1. GMT ratios suggested a superior antibody response with PHH-1V81 compared to BNT162b2 XBB.1.5 against XBB.1.5, while maintaining non-inferiority against JN.1. Noteworthy to mention that JN.1 VNA analysis was performed with a smaller subset of participants analysed and the GMT ratio was close to superiority. Moreover, upon PHH-1V81 vaccination, the RBD-responding IFN-γ-producing T-cells showed specificity not only against the homologous Omicron XBB.1.16 vaccine variant, but also cross-reactivity against the Omicron XBB.1.5, BA.1, BA.2.86 and JN.1 variants, and the ancestral Wuhan strain.

In subgroup analyses of neutralizing antibody responses against Omicron XBB.1.16 and Omicron XBB.1.5 among individuals aged 60 years or older, both those with and without prior reported SARS-CoV-2 infections, as well as subjects who received three or more prior doses, higher antibody titers were observed with PHH-1V81 compared to BNT162b2 XBB.1.5. While safety endpoint frequencies were similar between the two booster groups in all assessments, PHH-1V81 demonstrated an overall lower reactogenicity profile. Specifically, a significantly higher proportion of subjects in the PHH-1V81 arm reported no AE compared to those in the BNT162b2 XBB.1.5 arm. This trend aligns with the previously reported favorable safety profile of PHH-1V compared to BNT162b2.^18^ Moreover, it reinforces the notion that adjuvanted protein subunit vaccines, such as PHH-1V81, are particularly well-suited for vulnerable populations, including immunocompromised individuals, due to their safety profile and their ability to generate high levels of neutralizing antibodies, surpassing those induced by inactivated virus vaccines.^26^

The efficacy of the PHH-1V81 vaccine against Omicron’s XBB.1.16 and XBB.1.5 variants holds significant epidemiological importance amid Omicron’s emergence as the predominant SARS-CoV-2 variant globally.^27^ As evidenced by the emergence of the JN.1 variant, the current situation is characterized by the ongoing evolution of SARS-CoV-2 and the emergence of new variants with waning or poor protection from previous infections and vaccines as well as the potential ability to evade immunity and adapt for transmission.^6,28^ This issue is compounded by a population that has largely been previously exposed to infection with multiple variants, immunized and boosted.^29,30^ As a result, future vaccines may face challenges due to factors such as immune imprinting, immune seniority^31^ or an immunoglobulin G (IgG) class switch.^32^ In this complex scenario, it can be hypothesized that the squalene adjuvanted RBD-based vaccine can stimulate the innate immune system^33,34^ and, additionally, the RBD immune-focused approach can induce a better response against new and conserved epitopes in emerging variants,^35,36^ potentially overcoming or minimizing the potential negative effects associated with previous exposures. Furthermore, the IgG4 class switch associated with repeated vaccination with mRNA vaccines, suggests the potential for alternative platforms for subsequent immunizations.^32,37^ The results support these assumptions, as better responses were observed with PHH-1V81 compared to BNT162b2 XBB.1.5, against XBB and JN.1.

These interim results align with earlier findings of broad immune responses against previously circulating variants of concern (VOC), including Wuhan-Hu-1, Beta, Delta, and Omicron BA.1 observed with PHH-1V (Bimervax^®^; HIPRA).^18,38^ The ongoing phase IIb/III randomized controlled trial was adequately powered to assess the predetermined primary outcomes. The findings from this interim analysis underscore the strategic value of incorporating alternative vaccine platforms and adjuvants. This strategic shift aims to address the diminishing returns observed with successive mRNA vaccinations, ensuring sustained efficacy and immunogenicity against evolving variants of the virus.^39,40^ In this interim analysis, established laboratory techniques, including PBNA, VNA and ELISpot, were employed to evaluate immune responses. However, limitations stem from the interim nature of the results, the short follow-up period (14 days post-booster dose), limited statistical power for subgroup analyses, and a restricted representation of individuals aged 60 and older.

## 4. Conclusions

The interim findings strongly endorse the PHH-1V81 vaccine as a promising booster, offering balanced immunogenicity and tolerability. Such XBB updated vaccine is key for addressing the challenges posed by SARS-CoV-2 variability and population immune status, providing improved protection against current and emergent strains. Future research should prioritize prolonged follow-up to ascertain immune response persistence and effectiveness, especially among vulnerable groups, such as the immunosuppressed, those with chronic conditions, and elderly. With a favorable safety profile and robust response against the tested VOI, including the predominant JN.1, the adjuvanted RBD SARS-CoV-2 vaccine emerges as a compelling candidate for future COVID-19 vaccination strategies.

## Data Availability

All data relevant to the study are included in the article or uploaded as supplementary information. Further data are available from the authors upon reasonable request and with permission of HIPRA S.A.

## 5. Contributions

All authors had full access to all study data, interpreted the data, provided critical conceptual input, critically reviewed and revised the manuscript and approved the decision to submit for publication. MJLF, SN, AC, JMES, MJF, EAA, JM and SNM are PI from the participating hospitals and listed in order of subject contribution. JGP, RPC, LB and IE were involved specifically in cellular immune response assay development, data generation and data analysis. JB, NIU, DRR, RB were involved on humoral immune response assays development, data generation and data analysis. JPB was involved on data analysis and manuscript writing.

## 8. Acknowledgments

Special thanks to all the study participants who have voluntarily decided to participate in the study and without whom this would not have been possible, also to all health care professionals and researchers involved from participating hospitals and research institutionts.

Medical writing support was provided by Verónica Estévez and Silvia Paz (Smartworking4u S.L., Castellón, Spain) during the preparation of this paper and funded by HIPRA SCIENTIFIC, S.L.U.

Special thanks to all HIPRA members involved on study management, data management, pharmacovigilance, biostatistics analysis, quality, and thorough review of the manuscript.

We acknowledge the contribution of Ruth Peña for technical assistance with sample management and ELISpot and Felipe for data base generation

## Supplementary Appendix

Immunogenicity and safety of an Omicron XBB.1.16 adapted vaccine for COVID-19: Interim results from a randomized, controlled, multi-center clinical trial

### Inclusion and Exclusion Criteria

#### Inclusion Criteria

1. Participants must have met all the following criteria to be considered eligible for the study:
2. Adults aged 18 or older at Day 0.
3. Were willing and able to sign the informed consent and could comply with all study visits and procedures.
4. Participants must had received a primary scheme of an EU-approved mRNA vaccine (2 doses) and at least one booster dose with an EU-approved mRNA vaccine. Last booster dose must had been administered at least 6 months before Day 0.
5. Had a negative Rapid Antigen Test for COVID-19 at Day 0 prior to vaccination.
6. Adults determined by clinical assessment, including medical history and clinical judgement, to be eligible for the study, including adults with pre-existing chronic and stable diseases (non-immunocompromised), if these were stable and well-controlled according to the investigator’s judgement.
7. Participants biologically able to have children may have been enrolled in the study if the participant fulfilled all the following criteria:

- Had a negative urine pregnancy test at Day 0, only for those participants who were biologically able to become pregnant.
- Had practiced adequate contraception or had abstained from all activities that could result in pregnancy for at least 28 days prior to the study treatment, only for those participants who were biologically able to become pregnant.
- Had agreed to continue adequate contraception or abstinence through 3 months following the booster dose.

-Participants with female reproductive system:

i. Hormonal contraception [progesterone-only or combined: oral,
ii. injectable or transdermal (patch)]
iii. Intrauterine device
iv. Vasectomized partner (the vasectomized partner should be the sole
v. partner for that participant).
vi. Condom.
-Participants with male reproductive system:

i. Vasectomized participants.
ii. Agreed to use condom in partners biologically able to become
iii. pregnant.

#### Exclusion Criteria

Participants who met any of the following criteria were excluded from participation in this study:

1. Acute illness with fever ≥ 38.0°C at Day 0 or within 24 hours prior to vaccination. Afebrile participants with minor illnesses could be enrolled at the discretion of the investigator.
2. Other medical or psychiatric condition including recent (within the past year) or active suicidal ideation/behaviour that may have increased the risk of study participation or, in the investigator’s judgement, made the participant inappropriate for the study.

NOTE: This includes both conditions that may increase the risk associated with study intervention administration or a condition that may interfere with the interpretation of study results.
3. History of severe adverse reaction associated with a vaccine and/or severe allergic reaction (e.g., anaphylaxis) to any component of the study intervention.
4. Immunocompromised individuals defined as those with primary and secondary immune deficiencies and those receiving chemotherapy or immunosuppressant drugs other than steroids and glucocorticoids (maximum 30mg/day of prednisone, or equivalent, by any administration route for a maximum of 30 consecutive days), within 90 days prior to vaccination.
5. Bleeding diathesis or condition associated with prolonged bleeding that would, in the opinion of the investigator, contraindicate intramuscular injection.
6. Receipt of blood-derived immune globulins, blood, or blood-derived products in the past 3 months.
7. Participation in other studies involving study intervention if last dose was within 28 days prior to screening and/or it was planned to receive during study participation.
8. Received any non-study vaccine within 14 days before or after screening. For live or attenuated vaccines, 4 weeks before or after screening.
9. Received any COVID-19 vaccines other than EU-approved mRNA vaccines.
10. Received any Omicron XBB adapted vaccine before Day 0.
11. COVID-19 infection diagnosed in the previous 6 months before Day 0. History of COVID-19 infections was allowed.
12. History of a diagnosis or other conditions that, in the judgement of the investigator, may have affected study endpoint assessment or compromise participant safety.

**Figure S1.**
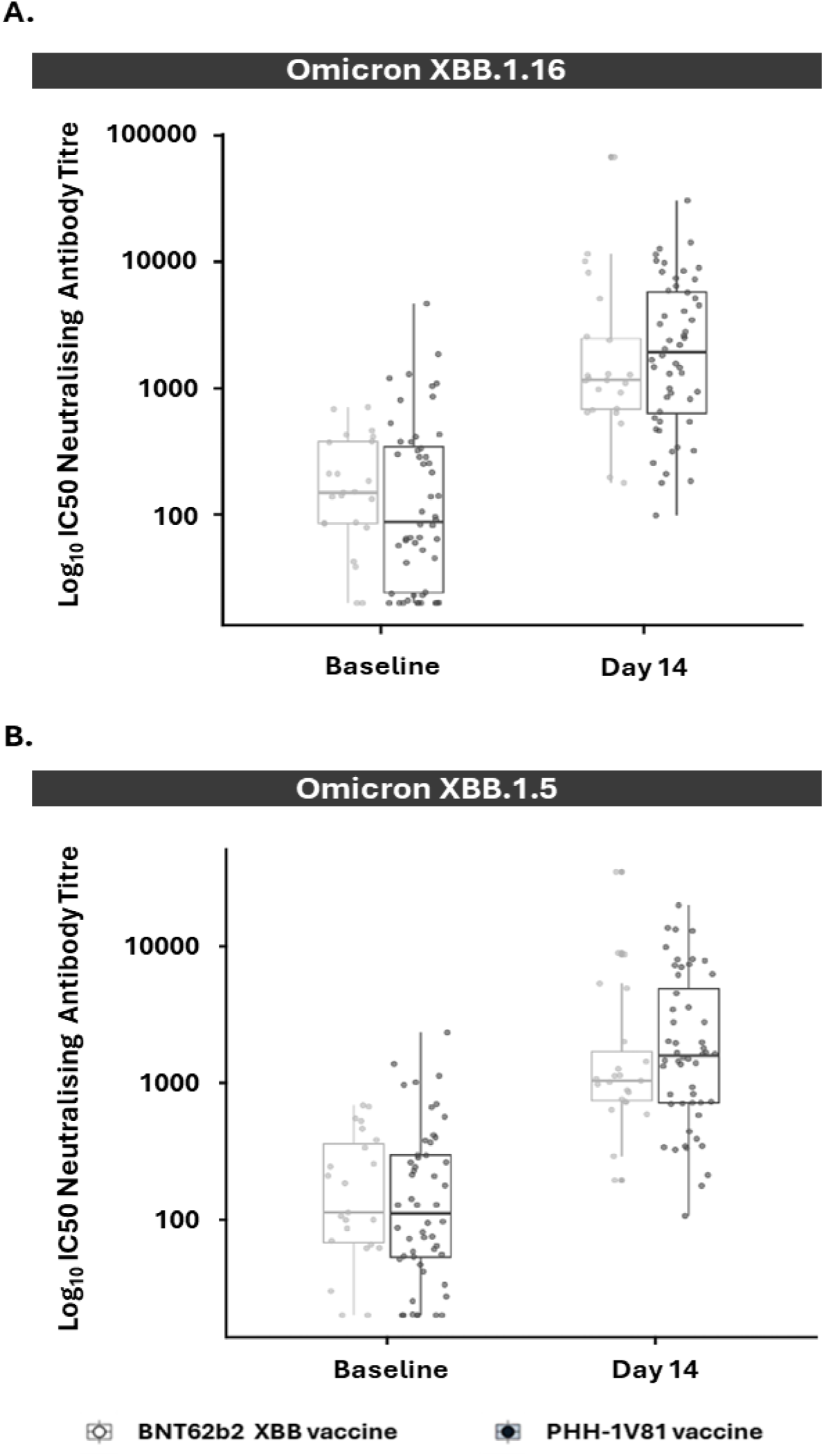
Neutralizing antibody responses against Omicron XBB.1.16 and Omicron XBB.1.5 variants in persons ≥60 years-old at baseline and day 14, by vaccine arm (mITT population) **A.**) Neutralizing antibody titre against Omicron XBB.1.16 variant on participants ≥ 60 years old (PHH-1V81, n=52; BNT62b2 XBB, n=23) for each vaccinated group; (**B.**) Neutralizing antibody titre against Omicron XBB.1.5 variant on participants ≥ 60 years old (PHH-1V81, n=52; BNT62b2 XBB, n=23) for each vaccinated group. Graphics represent individual log_10_ IC50 (dots) and box with median, IQR and whiskers of 1.5 times IQR. IC50: Half maximal inhibitory concentration; IQR: Interquartile range; SARS-CoV-2: severe acute respiratory syndrome coronavirus 2

**Figure S2.**
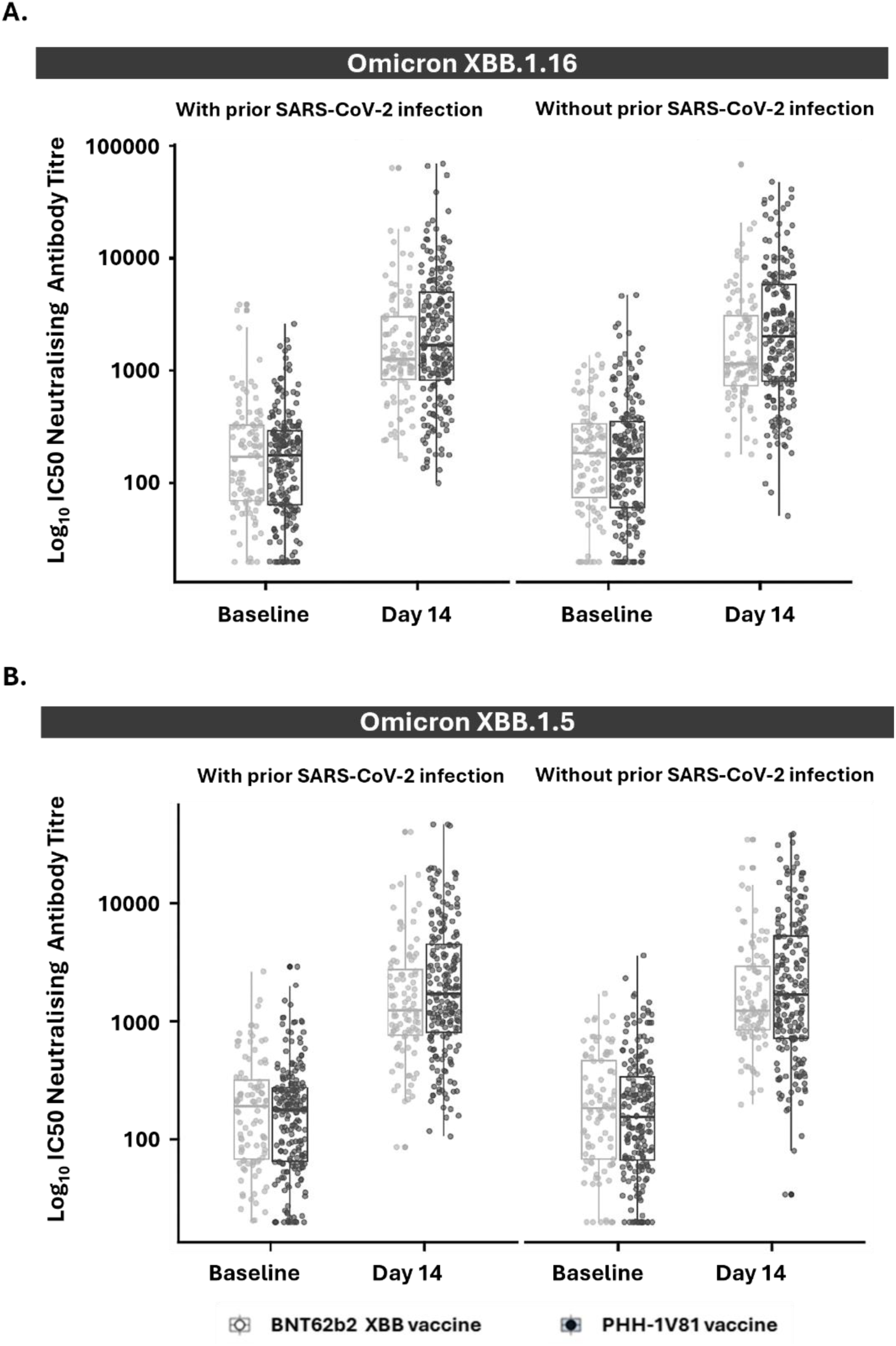
Neutralizing antibody responses against Omicron XBB.1.16 and Omicron XBB.1.5 variants in persons with and without prior reported SARS-CoV-2 infections at baseline and day 14, by vaccine arm (mITT population) **A.**) Neutralizing antibody titre against Omicron XBB.1.16 variant on participants with (PHH-1V81, n=206; BNT62b2 XBB, n=99) or without (PHH-1V81, n=200; BNT62b2 XBB, n=94) previous reported SARS-CoV-2 infection for each vaccinated group; (**B.**) Neutralizing antibody titre against Omicron XBB.1.5 variant on participants with (PHH-1V81, n=206; BNT62b2 XBB, n=99) or without (PHH-1V81, n=200; BNT62b2 XBB, n=94) previous reported SARS-CoV-2 infection for each vaccinated group. Graphics represent individual log_10_ IC50 (dots) and box with median, IQR and whiskers of 1.5 times IQR. IC50: Half maximal inhibitory concentration; IQR: Interquartile range; SARS-CoV-2: severe acute respiratory syndrome coronavirus 2

**Figure S3.**
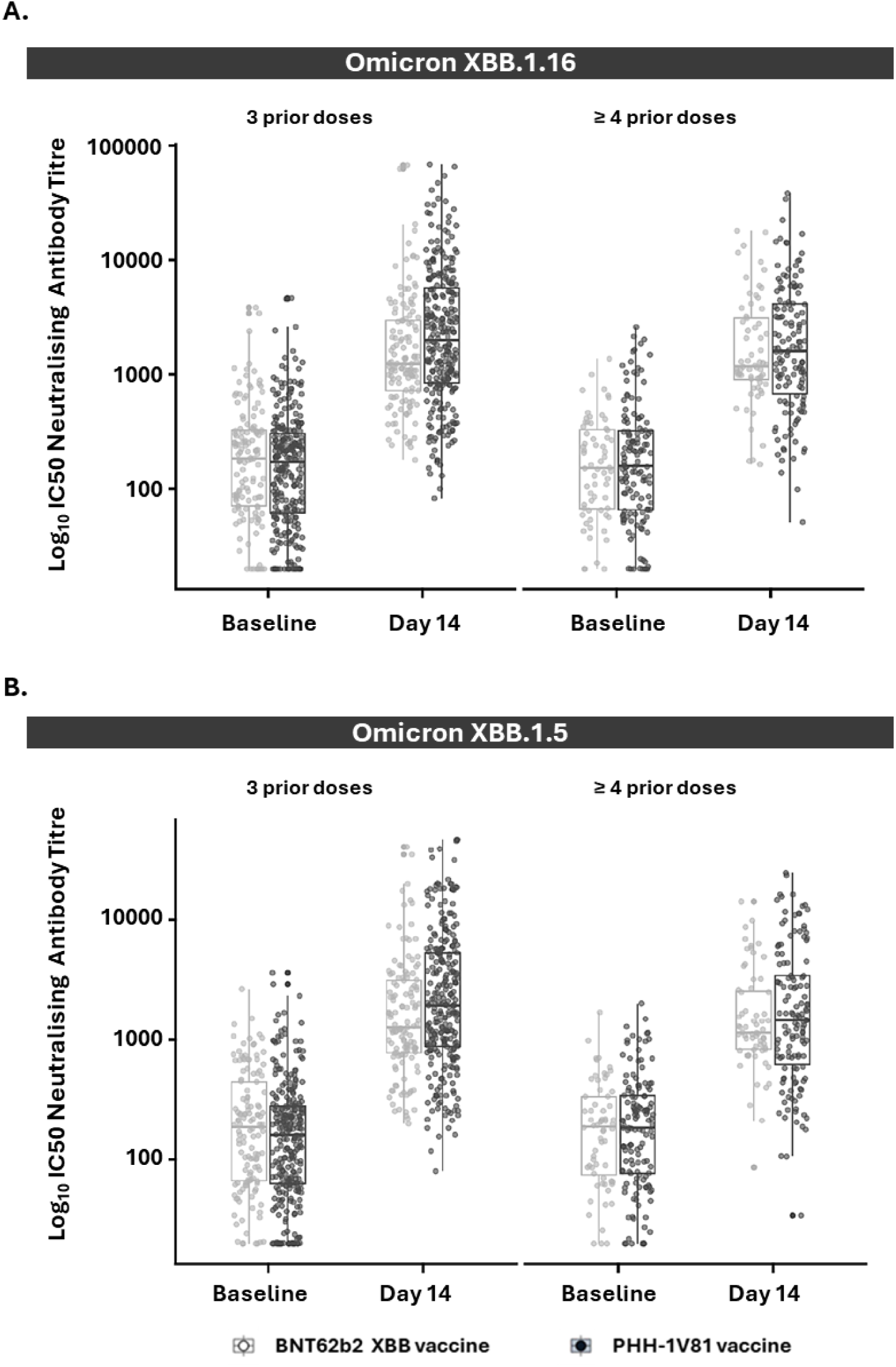
Neutralizing antibody responses against Omicron XBB.1.16 and Omicron XBB.1.5 variants in persons with 3 or ≥4 prior doses of a COVID-19 vaccine at baseline and day 14, by vaccine arm (mITT population) **A.**) Neutralizing antibody titre against Omicron XBB.1.16 variant on participants with 3 prior doses (PHH-1V81, n=272; BNT62b2 XBB, n=129) or ≥ 4 prior doses (PHH-1V81, n=134; BNT62b2 XBB, n=64) of SARS-CoV-2 vaccine for each vaccinated group; (**B.**) Neutralizing antibody titre against Omicron XBB.1.5 variant on participants with 3 prior doses (PHH-1V81, n=206; BNT62b2 XBB, n=99) or ≥ 4 prior doses (PHH-1V81, n=200; BNT62b2 XBB, n=94) of SARS-CoV-2 vaccine for each vaccinated group. Graphics represent individual log_10_ IC50 (dots) and box with median, IQR and whiskers of 1.5 times IQR. IC50: Half maximal inhibitory concentration; IQR: Interquartile range

**Table S1.** Baseline demographics of the safety population by vaccine arm.

|  | **PHH-1V81**<br>**(N=536)** | **BNT162b2 XBB.1.5**<br>**(N=264)** | **Total**<br>**(N=800)** |
| --- | --- | --- | --- |
| **Age** |  |  |  |
| n | 535 | 264 | 799 |
| Mean (SD; range), years | 44.7 (15.5; 18- 88) | 44.9 (15.0; 18- 86) | 44.7 (15.3; 18-88) |
| < 60 years old, n(%) | 463 (86.4%) | 233 (88.3%) | 696 (87.0%) |
| ≥60 years old, n(%) | 73 (13.6%) | 31 (11.7%) | 104 (13.0%) |
| **Gender** |  |  |  |
| Female, n(%) | 322 (60.1%) | 152 (57.6%) | 474 (59.3%) |
| **Race** |  |  |  |
| White, n(%) | 501 (93.5%) | 249 (94.3%) | 750 (93.8%) |
| Native Hawaiian or other Pacific Islander, n(%) | 1 (0.2%) | 0 (0.0%) | 1 (0.1%) |
| No reported, n(%) | 34 (6.3%) | 15 (5.7%) | 49 (6.1%) |
| **Prior reported COVID infection** |  |  |  |
| Yes | 278 (51.9%) | 134 (50.8%) | 412 (51.5%) |
| No | 258 (48.1%) | 130 (49.2%) | 388 (48.5%) |
| **Previous vaccine doses, n (%)** |  |  |  |
| 3 doses | 358 (66.8%) | 177 (67.0%) | 535 (66.9%) |
| 4 doses | 177 (33.0%) | 87 (33.0%) | 264 (33.0%) |
| 5 doses | 1 (0.2%) | 0 (0.0%) | 1 (0.1%) |
| N: the number of subjects in the population; n: the number of subjects meeting the criterion<br>SD: Standard deviation |  |  |  |

**Table S2.**
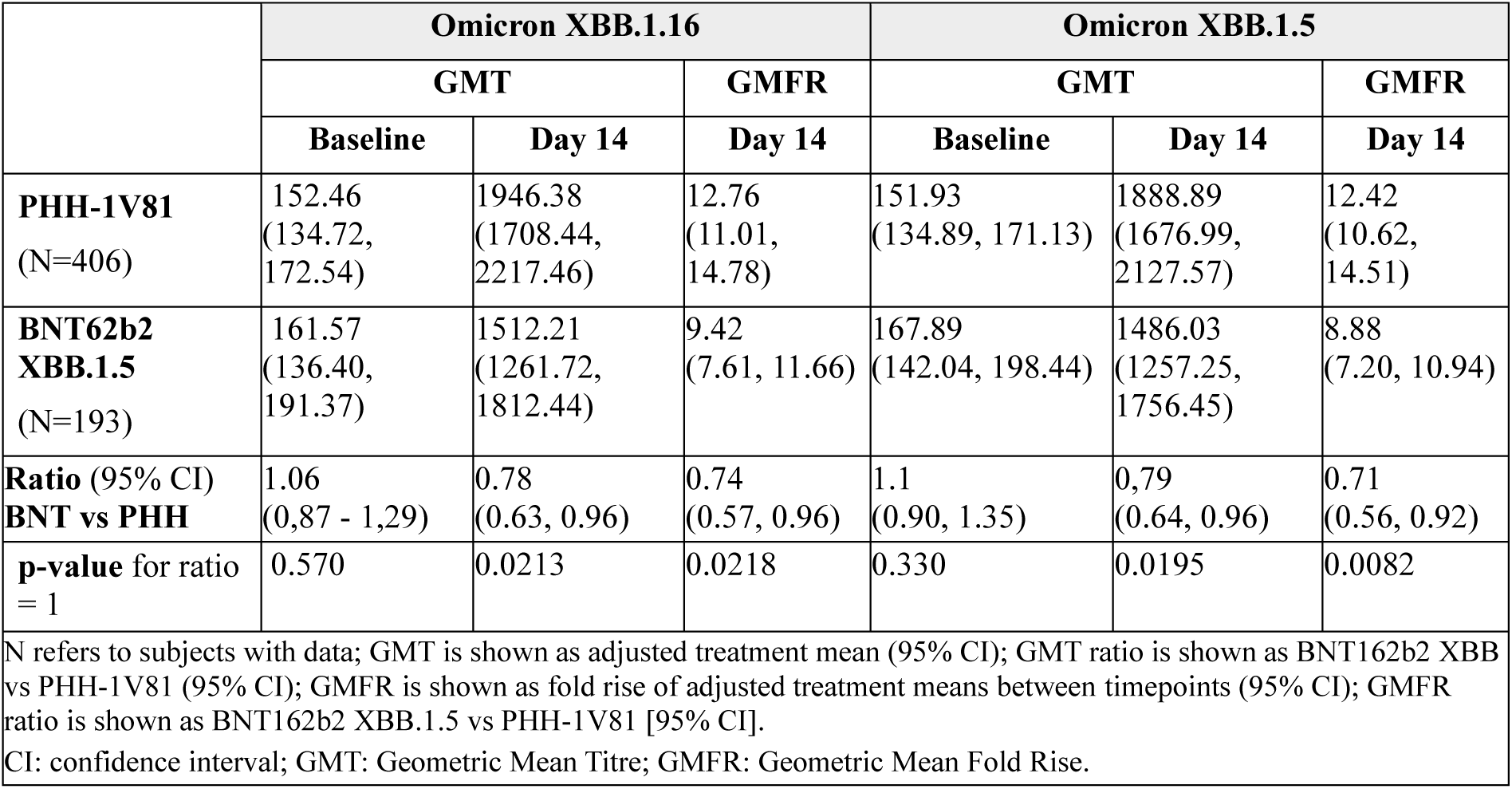
Analysis of neutralizing antibodies against Omicron XBB.1.16 and Omicron XBB.1.5 variants on Day 14 post-vaccination boost in the mITT population.

**Table S3.** Analysis of neutralizing antibodies against Omicron XBB.1.16 and Omicron XBB.1.5 variants in persons ≥60 years-old at Baseline and day 14 (mITT population)

| Variable | PHH-1V81<br>(N=52) | BNT162b2 XBB.1.5<br>(N=23) |
| --- | --- | --- |
| **Omicron XBB.1.16** |  |  |
| GMT (95% CI) at baseline | 127.94 (77.24; 211.91) | 174.81 (91.99; 332.19) |
| GMT (95% CI) at day 14 | 1979.2 (1194.91; 3278.25) | 1822.30 (958.92; 3463.01) |
| **Omicron XBB.1.5** |  |  |
| GMT (95% CI) at baseline | 136.61 (84.81; 220.05) | 173.46 (95.89; 313.80) |
| GMT (95% CI) at day 14 | 1817.99 (1128.63; 2928.39) | 1692.08 (935.35, 3061.04) |
| GMT is shown as adjusted treatment mean (95% CI)<br>CI: confidence interval; GMT: Geometric Mean Titre |  |  |

**Table S4.** Analysis of neutralizing antibodies against Omicron XBB.1.16 and Omicron XBB.1.5 variants in persons with and without prior reported SARS-CoV-2 infections at baseline and day 14 (mITT population), by vaccine arm (mITT population)

| Variable | With prior reported SARS-CoV-2 infection |  | Without prior reported SARS-CoV-2 infection |  |
| --- | --- | --- | --- | --- |
|  | PHH-1V81<br>(N=206) | BNT162b2<br>XBB.1.5<br>(N=99) | PHH-1V81<br>(N=200) | BNT162b2<br>XBB.1.5<br>(N=94) |
| **Omicron XBB.1.16** |  |  |  |  |
| GMT (95% CI) at baseline | 146.24<br>(123.06;<br>173.79) | 161.17<br>(127.18;<br>204.24) | 158.90<br>(130.95;<br>192.82) | 164.18<br>(126.1;<br>213.83) |
| GMT (95% CI) at day 14 | 1871.73<br>(1575.01;<br>2224.37) | 1493.63<br>(1178.65;<br>1892.78) | 2026.55<br>(1670.08;<br>2459.10) | 1562.60<br>(1199.81;<br>2035.07) |
| **Omicron XBB.1.5** |  |  |  |  |
| GMT (95% CI) at baseline | 148.38<br>(125.50;<br>175.43) | 166.05<br>(131.99;<br>208.91) | 154.17<br>(126.32;<br>188.15) | 171.63<br>(131.88;<br>223.35) |
| GMT (95% CI) at day 14 | 1880.24<br>(1590.31;<br>2223.01) | 1434.93<br>(1140.60;<br>1805.21) | 1879.39<br>(1539.94;<br>2293.68) | 1558.09<br>(1197.28;<br>2027.63) |
| GMT is shown as adjusted treatment mean (95% CI)<br>CI: confidence interval; GMT: Geometric Mean Titre; SARS-CoV-2: severe acute respiratory syndrome coronavirus 2 |  |  |  |  |

**Table S5.**
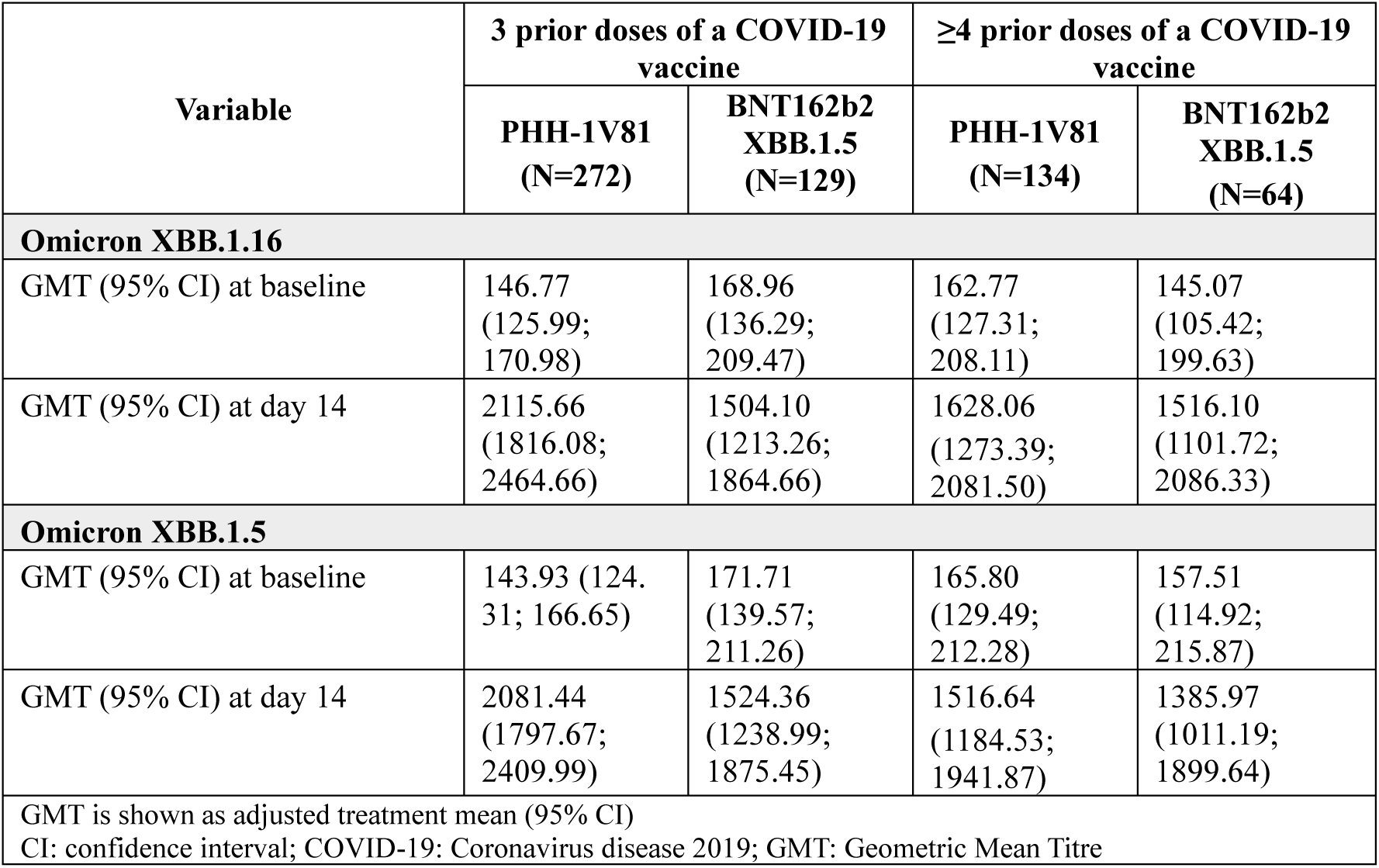
Analysis of neutralizing antibodies against Omicron XBB.1.16 and Omicron XBB.1.5 variants in persons with 3 or ≥4 prior doses of a COVID-19 vaccine at baseline and day 14, by vaccine arm (mITT population)

**Table S6.** Analysis of neutralizing antibodies against Omicron JN.1 variant by VNA.

|  | Omicron JN.1 by VNA |  |  |
| --- | --- | --- | --- |
|  | GMT |  | GMFR |
|  | Baseline | Day 14 | Day 14 |
| **PHH-1V81**<br>(N=65) | 58.51<br>(43.32, 79.02) | 768.44<br>(568.96, 1037.86) | 13.34<br>(8.84, 20.12) |
| **BNT162b2 XBB.1.5**<br>(N=35) | 53.02<br>(36.13, 77.82) | 505.88<br>(344.70, 742.43) | 9.27<br>(5.70, 15.07) |
| **Ratio (95% CI)**<br>**BNT vs PHH** | 0.91<br>(0.59, 1.39) | 0.66<br>(0.43, 1.01) | 0.69<br>(0.42, 1.14) |
| **p-value** for ratio = 1 | 0.6486 | 0.0540 | 0.1474 |
| N refers to subjects with data; GMT is shown as adjusted treatment mean (95% CI); GMT ratio is shown as BNT162b2 XBB vs PHH-1V81 (95% CI) followed by p-value for ratio = 1 ; GMFR is shown as fold rise of adjusted treatment means between timepoints (95% CI); GMFR ratio is shown as BNT162b2 XBB.1.5 vs PHH-1V81 [95% CI] followed by p-value for ratio = 1.<br>CI: confidence interval; GMT: Geometric Mean Titre; GMFR: Geometric Mean Fold Rise. |  |  |  |

**Table S7.** Analysis of IFN-γ producing T cells upon PBMC re-stimulation with SARS-CoV-2 derived peptide pools by ELISpot.

|  | PHH-1V81 | BNT162b2 XBB.1.5 |
| --- | --- | --- |
| **RBD Omicron XBB.1.16** |  |  |
| n | 27 | 12 |
| Median (Q1; Q3) at baseline | 33.75 (14.38; 64.38) | 31.25 (21.56; 57.19) |
| Median (Q1; Q3) at day 14 | 76.25 (24.38; 118.75) | 56.88 (163.44; 35.00) |
| Difference from baseline (SD) | 0.0029 (0.00093) | 0.0029 (0.00140) |
| p-value for difference = 0 | 0.0037 | 0.0454 |
| **RBD Omicron XBB.1.5** |  |  |
| n | 27 | 12 |
| Median (Q1; Q3) at baseline | 43.75 (16.25; 61.88) | 22.50 (14.06; 50.94) |
| Median (Q1; Q3) at day 14 | 60.00 (25.63; 128.13) | 53.13 (35.00; 133.13) |
| Difference from baseline (SD) | 0.0024 (0.00092) | 0.0033 (0.00137) |
| p-value for difference = 0 | 0.0139 | 0.0229 |
| **RBD Omicron JN.1** |  |  |
| n | 26 | 13 |
| Median (Q1; Q3) at baseline | 18.13 (7.71; 39.38) | 22.50 (8.75; 65.00) |
| Median (Q1; Q3) at day 14 | 36.88 (16.56; 66.56) | 32.50 (17.50; 61.25) |
| Difference from baseline (SD) | 0.0019 (0.00051) | 0.0015 (0.00072) |
| p-value for difference = 0 | 0.0007 | 0.0388 |
| Q1: lower quartile; Q3: upper quartile; SD: Standard deviation |  |  |

**Table S8.** Solicited systemic and local adverse events from day 0 through day 7 of the safety population that completed day 14 visit, by vaccine arm and type of adverse event.

| Type of events | PHH-1V81<br>(N=409) |  | BNT162b2 XBB.1.5<br>(N=198) |  | Overall<br>(N=607) |  |
| --- | --- | --- | --- | --- | --- | --- |
|  | Events | Subjects (%) | Events | Subjects (%) | Events | Subjects (%) |
| **Overall systemic events** | **282** | **114 (27.9)** | **155** | **65 (32.8)** | **437** | **179 (29.5)** |
| Headache | 80 | 71 (17.4) | 45 | 40 (20.2) | 125 | 111 (18.3) |
| Fatigue | 60 | 56 (13.7) | 35 | 32 (16.2) | 95 | 88 (14.5) |
| Muscle Pain | 54 | 48 (11.7) | 25 | 24 (12.1) | 79 | 72 (11.9) |
| Malaise/Discomfort | 36 | 33 (8.1) | 25 | 24 (12.1) | 61 | 57 (9.4) |
| Diarrhoea | 16 | 15 (3.7) | 6 | 6 (3.0) | 22 | 21 (3.5) |
| Nausea/ Vomiting | 14 | 13 (3.2) | 5 | 5 (2.5) | 19 | 18 (3.0) |
| Enlarged lymphnodes<br>(lymphadenopathy) | 11 | 11 (2.7) | 6 | 6 (3.0) | 17 | 17 (2.8) |
| Axillary pain | 9 | 9 (2.2) | 7 | 7 (3.5) | 16 | 16 (2.6) |
| Fever | 2 | 2 (0.5) | 1 | 1 (0.5) | 3 | 3 (0.5) |
| **Overall local events** | **502** | **214 (52.3)** | **284** | **120 (60.6)** | **786** | **334 (55.0)** |
| Injection site pain / tenderness/<br>discomfort | 362 | 210 (51.3) | 201 | 117 (59.1) | 563 | 327 (53.9) |
| Injection site induration / swelling | 85 | 65 (15.9) | 58 | 42 (21.2) | 143 | 107 (17.6) |
| Injection site erythema / redness | 55 | 46 (11.2) | 25 | 22 (11.1) | 80 | 68 (11.2) |

